# The Association of Marital/Partner Status with Patient-Reported Health Outcomes Following Acute Myocardial Infarction or Stroke: Protocol for a Systematic Review and Meta-analysis

**DOI:** 10.1101/2022.04.26.22273986

**Authors:** Cenjing Zhu, Phoebe M Tran, Erica C Leifheit, Erica S Spatz, Rachel P Dreyer, Kate Nyhan, Shi-Yi Wang, Larry B Goldstein, Judith H Lichtman

**Author notes:** Corresponding author Address: 60 College Street, New Haven, 06511.

## Abstract

**Introduction:** Marital/Partner support is associated with lower mortality and morbidity following acute myocardial infarction (AMI) and stroke. Despite an increasing focus on the effect of patient-centered factors on health outcomes, little is known about the impact of marital/partner status on patient-reported outcome measures (PROMs).

**Objective:** To synthesize evidence of the association between marital/partner status and PROMs after AMI and stroke and to determine whether associations differ by sex.

**Methods and analysis:** We will search MEDLINE (via Ovid), Web of Science Core Collection (as licensed by Yale University), Scopus, EMBASE (via Ovid), and PsycINFO (via Ovid) from inception to January 10, 2022. Two authors will independently screen titles, abstracts, and then full texts as appropriate, extract data, and assess risk of bias. Conflicts will be resolved by discussion with a third reviewer. The primary outcomes will be the associations between marital/partner status and PROMs. Meta-analysis will be conducted if appropriate. Subgroup analysis by sex and meta-regression with a covariate for the proportion of male participants will be performed to explore differences by sex.

**Ethics and dissemination:** This research is exempt from ethics approval because the study will be conducted using published data. We will disseminate the results of the analysis in a related peer-reviewed journal.

**PROSPERO registration number:** CRD42022295975

## Introduction

Marital/Partner status is an important social factor that affects acute myocardial infarction (AMI) and stroke outcomes. Being married/partnered has been associated with lower mortality and higher event-free survival following an AMI or stroke, supported by studies varying in design, setting, and scale.(1–6) However, little is known about the impact of marital/partner status on patient-centered outcomes.

Patient-reported outcome measures (PROMs) are defined by the Food & Drug Administration and National Quality Forum as outcomes that derive directly from the patient about the status of a patient’s health condition without amendment or interpretation of the patient’s response by a clinician.(7,8) With strides in cardiovascular disease and stroke, numerous PROMs have been developed, validated, and used in clinical studies to quantify treatment benefits with regard to improvements in symptoms, functional outcomes, and health-related quality of life.(9–11) These measures also independently predict subsequent cardiovascular events, hospitalizations, costs of care, and mortality, and they have the potential to inform clinical decision making and targets for risk adjustment.(12)

Prior studies assessed patient-reported health status outcomes among AMI and stroke patients using different PROMs, but the results regarding the association between marital/partner status and outcomes are inconsistent.(13–16) No systematic review investigates and synthesizes the association of marital/partner status with PROMs among individuals who have had an AMI or stroke. Further, although women may not benefit from marriage to the same extent as men regarding outcomes such as mortality and major adverse cardiac events,(2,17,18) less is known about whether there are sex differences in the degree of “protection” conferred by marriage or partnership during AMI and stroke recovery as assessed with PROMs.

## Objective

This purpose of this report is to establish the explicit methodology we will use for conducting a systematic review and meta-analysis to answer the following questions: 1, does marital/partner status impact the patient-reported health status outcomes of individuals who have had an AMI or stroke; and 2, are there sex differences in the impact of marital/partner status on patient-reported health status outcomes of individuals who had an AMI or stroke?

## Methods

The review protocol will comply with the Preferred Reporting Items for Systematic Reviews and Meta-Analyses Protocols (PRISMA-P) statement guidance (19) for reporting the present protocol and subsequent formal meta-analysis (**Supplement Table 1**). The review protocol is registered on the International Prospective Register of Systematic Reviews (PROSPERO) database (ID: CRD42022295975).

### Search strategy and information sources

We will conduct a literature search using Medline (via Ovid), Web of Science Core Collection (as licensed by Yale University), Scopus, EMBASE (via Ovid), and PsycINFO (via Ovid) to identify publications from inception to January 10, 2022. For each concept in the search strategy, we will use appropriate keywords and subject indexing terms, informed by the search strategies used in previous reports. The search strategy was developed and adapted for individual databases and interfaces as needed (details in **Supplement Table 2**). In addition, ProQuest Theses and Dissertations, OpenGrey, and ClinicalTrials.gov will be searched for dissertations and grey literature. For each included article that meets the eligibility criteria, we will conduct backward and forward citation chaining in The Lens via Citation Chaser.(20)

### Eligibility criteria

The systematic review will identify studies that evaluate marital/partner status as an independent variable and report their associations with one or more defined PROMs. The included studies should have at least two groups (married vs. unmarried, or partnered vs. not partnered). Studies with no clearly indicated reference/control group or defined outcome will be excluded. Study participants will include individuals aged 18 years and older who were diagnosed with an AMI or stroke by a medical professional. PROMs may be obtained from proxy respondents if the patient was not able to answer. Animal studies will not be included. No restrictions will be placed on study type, setting, time frame, or publication year. Only reports that are available in print or downloadable form and written in English will be included.

### Data extraction and management

Records identified from the database search will be uploaded into a Covidence project. After deduplication, unique records will be screened independently at the title-abstract stage by two reviewers (C.Z. and P.T.) using pre-specified inclusion criteria. The full text of the identified studies will then be assessed by the two reviewers, with documentation of the reasons for any exclusions. A third researcher will help the screeners achieve consensus in cases of disagreement.

After screening, two researchers will independently extract the data on author information, publication year, country, study type (observational vs. interventional), study setting (institutionalized vs. community-dwelling), features of the study participants, the main exposure definition(s) (married/partnered vs. unmarried/not partnered or divorced/separated, widowed), the main outcome definition(s) (PROM, name, type, measurement domain, time points), the number of individuals with the outcome based on the exposure, the effect estimates (unadjusted and/or adjusted odds ratios, risks ratios, hazard ratios, or regression coefficients [β] with standard errors [SEs], which can be calculated from 95% confidence intervals [CIs] or p-values) and adjustment factors, sex-specific estimates (if reported), and disease type (AMI or stroke).

The primary outcome of the review is the association between marital/partner status and PROMs. Relevant data will be recorded using a standardized data extraction form (**Supplement Table 3**), which may be adapted during the extraction process to capture additional information if needed. Authors of the included studies will be contacted for missing key information.

### Quality assessment

Methodological quality (risk of bias) of the included studies will be appraised using the Newcastle-Ottawa Scale.(21,22) This scale is the most commonly used methodological quality assessment measure for observational studies. The scale output will be converted to the Agency for Healthcare Research and Quality standard (good, fair, poor quality) based on previously defined thresholds.(22)

### Data synthesis and subgroup analysis

After data extraction is completed, we will determine the feasibility of conducting a meta-analysis for each PROM (≥2 studies reporting effect measures and 95% CIs for a PROM) and whether meta-analysis is appropriate (i.e., if the features of these studies are sufficiently similar to combine). If conducting a meta-analysis is feasible and appropriate, the extracted effect estimates with SEs will be combined using a random-effects model with inverse-variance weighting to calculate pooled effect sizes. For studies reporting both unadjusted and adjusted effect estimates, the adjusted effect judged to minimize the risk of bias due to confounding will be used for the meta-analysis.(23) The heterogeneity of the studies will be measured with I^2^ statistics (I^2^ values of <25%, 25%-50%, 51%-75%, □and >75% indicate low, moderate, medium, and high heterogeneity, respectively), which describe the variation of effect size that is attributable to heterogeneity across studies.(24) Visual assessment of the funnel plot and Egger’s test will be used to assess for publication bias.(25) If a meta-analysis is not feasible or appropriate, we will instead conduct a narrative synthesis of the studies. The Grading of Recommendations Assessment, Development, and Evaluation (GRADE) system (26) will be used to evaluate the strength of the evidence body.

Separate analysis will be conducted by type of disease (AMI or stroke) and subgroup analyses will be performed according to sex. To investigate the sources of heterogeneity, meta-regression will be performed adjusting for the covariates, including the duration of follow-up, study setting (hospital-based vs. population-based), bias score, mean age, male proportion, and sample size. If there is high heterogeneity, we will further conduct subgroup analyses according to the significant covariates. Sensitivity analysis will be performed by excluding studies with “low quality” according to the Newcastle-Ottawa Scale and comparing the results with the overall analysis.

### Ethics and dissemination

This study is exempt from institutional review board approval because it is a secondary analysis of published or aggregated data. No new patients will be involved in this study. The data collected during the review will be made fully available without restriction upon study completion.

## Discussion

AMI and stroke represent a considerable population disease burden and are leading causes of death and disability worldwide.(27,28) Although advances in prevention, diagnosis, and therapeutics led to increasing numbers of people surviving AMI and stroke, these diseases impact many aspects of the lives of affected individuals following hospital discharge.(28)

A better prognosis after AMI and stroke among those who are married or have partners is supported by studies varying in design, setting, and scale.(1–6) A meta-analysis of 34 studies found a 42% higher mortality for unmarried AMI patients compared to their married counterparts.(1) Another review provided evidence on the association of marriage with longer event-free survival and better risk factor control among cardiovascular patients.(4) At least one study reported poorer outcomes after stroke for those who were not married.(29) Despite existing review articles focusing on mortality and morbidity outcomes, there remains a lack of understanding of the impact of marital/partner status on patient-centered outcomes measured by PROMs.

Observational studies of patients with AMI and stroke assessing the associations between marital/partner status and patient-reported health status outcomes varied in study population, PROMs used to measure health status, and period of data collection relative to the index event. The results of these studies are inconsistent. For example, three studies found that marital status was associated with quality of life among those who had an AMI or stroke,(13,14,16) but another found no association.(15) By systematically reviewing the literature and assessing the available evidence, we aim to clarify and quantify the association of marital/partner status with patient-centered outcomes measured using PROMs and explore differences by sex. A better understanding of this gap in knowledge can inform the design of secondary prevention interventions to improve long-term recovery of those who had an AMI or stroke, and potentially support targeted interventions.

This systematic review will face several challenges. First, although studies with proxy responses are eligible for inclusion in the review, patients who have specific stroke-related deficits such as aphasia or cognitive impairments may be excluded from some studies according to their prespecified criteria, introducing potential bias. Second, we will conduct our literature search using multiple databases, but articles published in languages other than English will not be eligible. Third, because we will consider a variety of study designs and settings, heterogeneity between studies is anticipated to be high. We plan to extract a set of study characteristics and conduct meta-regression to investigate the sources of heterogeneity. Subgroup analysis will be performed based on the significant covariates identified from the meta-regression.

## Data Availability

No datasets were generated or analyzed during the current study. All relevant data from this study will be made available upon study completion.

## Authors’ contributions

Conceptualization (C.Z., P.T., J.L.), formal analysis (C.Z., P.T., K.N), Methodology (C.Z., K.N., S.W., J.L.), Writing – Original Draft Preparation (C.Z.), Writing – Review & Editing (P.T., E.S., E.L., R.D., K.N., S.W., L.G., J.L.)

## Acknowledgement

None

## Notes

### Competing Interest Statement

The authors have declared no competing interest.

### Funding Statement

The author(s) received no specific funding for this work.

## References

1. Wong CW, Kwok CS, Narain A, Gulati M, Mihalidou AS, Wu P, et al. Marital status and risk of cardiovascular diseases: a systematic review and meta-analysis. Heart. 2018 Dec;104(23):1937–48.

2. Wang Y, Jiao Y, Nie J, O’Neil A, Huang W, Zhang L, et al. Sex differences in the association between marital status and the risk of cardiovascular, cancer, and all-cause mortality: a systematic review and meta-analysis of 7,881,040 individuals. Glob Health Res Policy. 2020 Feb 28;5:4.

3. Dhindsa DS, Khambhati J, Schultz WM, Tahhan AS, Quyyumi AA. Marital status and outcomes in patients with cardiovascular disease. Trends in Cardiovascular Medicine. 2020 May 1;30(4):215–20.

4. Manfredini R, De Giorgi A, Tiseo R, Boari B, Cappadona R, Salmi R, et al. Marital Status, Cardiovascular Diseases, and Cardiovascular Risk Factors: A Review of the Evidence. J Womens Health (Larchmt). 2017 Jun;26(6):624–32.

5. Damiani G, Salvatori E, Silvestrini G, Ivanova I, Bojovic L, Iodice L, et al. Influence of socioeconomic factors on hospital readmissions for heart failure and acute myocardial infarction in patients 65 years and older: evidence from a systematic review. Clin Interv Aging. 2015 Jan 12;10:237–45.

6. Andersen KK, Olsen TS. Stroke case-fatality and marital status. Acta Neurol Scand. 2018 Oct;138(4):377–83.

7. NQF: Patient-Reported Outcomes [Internet]. [cited 2021 Dec 20]. Available from: https://www.qualityforum.org/Projects/n-r/Patient-Reported_Outcomes/Patient-Reported_Outcomes.aspx

8. Guidance for Industry on Patient-Reported Outcome Measures: Use in Medical Product Development to Support Labeling Claims; Availability [Internet]. Federal Register. 2009 [cited 2021 Dec 20]. Available from: https://www.federalregister.gov/documents/2009/12/09/E9-29273/guidance-for-industry-on-patient-reported-outcome-measures-use-in-medical-product-development-to

9. Rumsfeld JS, Alexander KP, Goff DC, Graham MM, Ho PM, Masoudi FA, et al. Cardiovascular Health: The Importance of Measuring Patient-Reported Health Status. Circulation. 2013 Jun 4;127(22):2233–49.

10. Scoggins JF, Patrick DL. The use of patient-reported outcomes instruments in registered clinical trials: Evidence from ClinicalTrials.gov. Contemporary Clinical Trials. 2009 Jul 1;30(4):289–92.

11. Reeves M, Lisabeth L, Williams L, Katzan I, Kapral M, Deutsch A, et al. Patient-Reported Outcome Measures (PROMs) for Acute Stroke: Rationale, Methods and Future Directions. Stroke. 2018 Jun 1;49(6):1549–56.

12. Mommersteeg PMC, Denollet J, Spertus JA, Pedersen SS. Health status as a risk factor in cardiovascular disease: A systematic review of current evidence. American Heart Journal. 2009 Feb 1;157(2):208–18.

13. Wlodarczyk D, Zietalewicz U. How gender-specific are predictors of post-MI HRQoL? A longitudinal study. Health Qual Life Outcomes. 2020 Jun 26;18:202.

14. Lisiak M, Uchmanowicz I, Wontor R. Frailty and quality of life in elderly patients with acute coronary syndrome. CIA. 2016 May 5;11:553–62.

15. Abualait TS, Alzahrani MA, Ibrahim AI, Bashir S, Abuoliat ZA. Determinants of life satisfaction among stroke survivors 1 year post stroke. Medicine (Baltimore). 2021 Apr 23;100(16):e25550.

16. Rachpukdee S, Howteerakul N, Suwannapong N, Tang-aroonsin S. Quality of Life of Stroke Survivors: A 3-Month Follow-up Study. Journal of Stroke and Cerebrovascular Diseases. 2013 Oct 1;22(7):e70–8.

17. Robards J, Evandrou M, Falkingham J, Vlachantoni A. Marital status, health and mortality. Maturitas. 2012 Dec;73(4):295–9.

18. Manzoli L, Villari P M Pirone G, Boccia A. Marital status and mortality in the elderly: A systematic review and meta-analysis. Social Science & Medicine. 2007 Jan 1;64(1):77–94.

19. Moher D, Shamseer L, Clarke M, Ghersi D, Liberati A, Petticrew M, et al. Preferred reporting items for systematic review and meta-analysis protocols (PRISMA-P) 2015 statement. Syst Rev. 2015 Jan 1;4(1):1.

20. Haddaway NR, Grainger MJ, Gray CT. citationchaser: An R package and Shiny app for forward and backward citations chasing in academic searching [Internet]. Zenodo; 2021 [cited 2021 Dec 7]. Available from: https://zenodo.org/record/4543513

21. Ma L-L, Wang Y-Y, Yang Z-H, Huang D, Weng H, Zeng X-T. Methodological quality (risk of bias) assessment tools for primary and secondary medical studies: what are they and which is better? Mil Med Res. 2020 Feb 29;7:7.

22. Wells G, Shea B, O’Connell D, Peterson J, Welch V, Losos M, et al. The Newcastle-Ottawa scale (NOS) for assessing the quality of nonrandomised studies in meta-analyses. [Internet]. 2016 [cited 2021 Dec 21]. Available from: http://www.ohri.ca/programs/clinical_epidemiology/oxford.asp

23. Chapter 24: Including non-randomized studies on intervention effects [Internet]. [cited 2022 Feb 15]. Available from: https://training.cochrane.org/handbook/current/chapter-24

24. Melsen WG, Bootsma MCJ, Rovers MM, Bonten MJM. The effects of clinical and statistical heterogeneity on the predictive values of results from meta-analyses. Clinical Microbiology and Infection. 2014 Feb 1;20(2):123–9.

25. Sterne JAC, Egger M, Smith GD. Investigating and dealing with publication and other biases in meta-analysis. BMJ. 2001 Jul 14;323(7304):101–5.

26. Owens DK, Lohr KN, Atkins D, Treadwell JR, Reston JT, Bass EB, et al. AHRQ Series Paper 5: Grading the strength of a body of evidence when comparing medical interventions—Agency for Healthcare Research and Quality and the Effective Health-Care Program. Journal of Clinical Epidemiology. 2010 May 1;63(5):513–23.

27. Rudd KE, Johnson SC, Agesa KM, Shackelford KA, Tsoi D, Kievlan DR, et al. Global, regional, and national sepsis incidence and mortality, 1990–2017: analysis for the Global Burden of Disease Study. Lancet. 2020 Jan 18;395(10219):200–11.

28. Virani SS, Alonso A, Aparicio HJ, Benjamin EJ, Bittencourt MS, Callaway CW, et al. Heart Disease and Stroke Statistics—2021 Update. Circulation. 2021 Feb 23;143(8):e254–743.

29. Liu Q, Wang X, Wang Y, Wang C, Zhao X, Liu L, et al. Association between marriage and outcomes in patients with acute ischemic stroke. J Neurol. 2018;265(4):942–8.

