## Supplemental tables for "The Association of Marital/Partner Status with Patient-Reported Health Outcomes Following Acute Myocardial Infarction or Stroke: Protocol for a Systematic Review and Meta-analysis"

**Supporting information**

Supplemental Table 1. PRISMA-P (Preferred Reporting Items for Systematic review and Meta-Analysis Protocols) 2015 checklist

Supplemental Table 2. Search strategy

Supplemental Table 3. Data extraction form

**Supplement Table 1. PRISMA-P (Preferred Reporting Items for Systematic review and Meta-Analysis Protocols) 2015 checklist: recommended items to address in a systematic review protocol**

| Section and topic | Item No | Checklist item | Considered |
| --- | --- | --- | --- |
| ADMINISTRATIVE INFORMATION | | |  |
| Title: |  |  |  |
| Identification | 1a | Identify the report as a protocol of a systematic review | Page 1 |
| Update | 1b | If the protocol is for an update of a previous systematic review, identify as such | NA |
| Registration | 2 | If registered, provide the name of the registry (such as PROSPERO) and registration number | Page 1 |
| Authors: |  |  |  |
| Contact | 3a | Provide name, institutional affiliation, e-mail address of all protocol authors; provide physical mailing address of corresponding author | Page 1 |
| Contributions | 3b | Describe contributions of protocol authors and identify the guarantor of the review | Page 7 |
| Amendments | 4 | If the protocol represents an amendment of a previously completed or published protocol, identify as such and list changes; otherwise, state plan for documenting important protocol amendments | NA |
| Support: |  |  | NA |
| Sources | 5a | Indicate sources of financial or other support for the review |  |
| Sponsor | 5b | Provide name for the review funder and/or sponsor |  |
| Role of sponsor or funder | 5c | Describe roles of funder(s), sponsor(s), and/or institution(s), if any, in developing the protocol |  |
| INTRODUCTION | | |  |
| Rationale | 6 | Describe the rationale for the review in the context of what is already known | Page 3 |
| Objectives | 7 | Provide an explicit statement of the question(s) the review will address with reference to participants, interventions, comparators, and outcomes (PICO) | Page 3-4 |
| METHODS | | |  |
| Eligibility criteria | 8 | Specify the study characteristics (such as PICO, study design, setting, time frame) and report characteristics (such as years considered, language, publication status) to be used as criteria for eligibility for the review | Page 4 |
| Information sources | 9 | Describe all intended information sources (such as electronic databases, contact with study authors, trial registers or other grey literature sources) with planned dates of coverage | Page 4 |
| Search strategy | 10 | Present draft of search strategy to be used for at least one electronic database, including planned limits, such that it could be repeated | Page 4-5, S2 |
| Study records: |  |  |  |
| Data management | 11a | Describe the mechanism(s) that will be used to manage records and data throughout the review | Page 5 |
| Selection process | 11b | State the process that will be used for selecting studies (such as two independent reviewers) through each phase of the review (that is, screening, eligibility and inclusion in meta-analysis) | Page 4-5 |
| Data collection process | 11c | Describe planned method of extracting data from reports (such as piloting forms, done independently, in duplicate), any processes for obtaining and confirming data from investigators | Page 5 |
| Data items | 12 | List and define all variables for which data will be sought (such as PICO items, funding sources), any pre-planned data assumptions and simplifications | Page 5 |
| Outcomes and prioritization | 13 | List and define all outcomes for which data will be sought, including prioritization of main and additional outcomes, with rationale | Page 5 |
| Risk of bias in individual studies | 14 | Describe anticipated methods for assessing risk of bias of individual studies, including whether this will be done at the outcome or study level, or both; state how this information will be used in data synthesis | Page 5 |
| Data synthesis | 15a | Describe criteria under which study data will be quantitatively synthesized | Page 5 |
|  | 15b | If data are appropriate for quantitative synthesis, describe planned summary measures, methods of handling data and methods of combining data from studies, including any planned exploration of consistency (such as I^2^, Kendall’s τ) | Page 5-6 |
|  | 15c | Describe any proposed additional analyses (such as sensitivity or subgroup analyses, meta-regression) | Page 6 |
|  | 15d | If quantitative synthesis is not appropriate, describe the type of summary planned | Page 5-6 |
| Meta-bias(es) | 16 | Specify any planned assessment of meta-bias(es) (such as publication bias across studies, selective reporting within studies) | Page 6 |
| Confidence in cumulative evidence | 17 | Describe how the strength of the body of evidence will be assessed (such as GRADE) | Page 6 |

*NA: Not applicable.

*From: Shamseer L, Moher D, Clarke M, Ghersi D, Liberati A, Petticrew M, Shekelle P, Stewart L, PRISMA-P Group. Preferred reporting items for systematic review and meta-analysis protocols (PRISMA-P) 2015: elaboration and explanation. BMJ. 2015 Jan 2;349(jan02 1):g7647.*

**Supplement Table 2. Search strategy**

|  | **MeSH** | **Keywords** | **Notes** |
| --- | --- | --- | --- |
| **Exposure:**  **Marital/Partner status** | marital status  (narrower terms: “divorce”, “marriage”, “spouses”, “marital status”,  “single person”,  “widowhood”) | marital/spousal relationship, unmarried, **partnership**, marriage, wife/husband, spouse, **domestic partner**, break-up, divorce*, widow*, single, spinster, bachelor, girlfriend, boyfriend | We will only consider papers that assess marital/relationship status as an independent exposure. We will not consider a composite exposure that includes marital status (e.g., ‘cohabitation’ if defined based on both marital status and living condition; or living with spouse or family members). |
| **Disease:**  **AMI or stroke** | myocardial infarction, acute coronary syndrome, coronary disease | coronary heart disease, ischemic/ischaemic heart disease, ischemic/ischaemic heart disease, heart attack, AMI, |  |
|  | stroke  cerebrovascular disorders | cerebrovascular disease, cerebrovascular accident |  |
| **Outcome:**  **Patient-reported outcome measures** | patient reported outcome measures (2017); patient outcome assessment (2013-2016) | patient-reported outcome*, self-report*, patient-reported*, patient-centered*, preference, experience, perception, perceived, measure*, questionnaire*, survey*, psychometric*, depress*, psychologic*, anxiety, symptom*, fatigue | **3 approaches:**  1. Oxford filter for PROM: <https://cosmin.nl/wp-content/uploads/prom-search-filter-oxford-2010.pdf> (available for Ovid and PubMed, translated to Web of Science and Scopus by C.Z.)  2. MeSH ("Patient Reported Outcome Measures"; "Patient Outcome Assessment") + keywords search for “patient-reported health status”  3. Identified from literature, scoping search (via [ResearchRabbit](https://researchrabbitapp.com/home)) and [NINDS CDE](https://www.commondataelements.ninds.nih.gov/Stroke), most commonly used PROMs in AMI/stroke:   - Generic: EQ-5D, Short Form-12/36, General Health Questionnaire, PROMIS - Stroke-specific: Neuro-QOL, Stroke Impact Scale, Stroke Specific QOL, SAQOL, SA-SIP - AMI-specific: QLMI, Seattle Angina Questionnaire, Mac-New Questionnaire, HeartQoL, QLICD_CHD |

**2.1 Search strategy for MEDLINE (via Ovid):**

<https://ovidsp.ovid.com/ovidweb.cgi?T=JS&NEWS=N&PAGE=main&SHAREDSEARCHID=6gNZbHQgMSBhTX2kpLQvHgddI1nPVe56a09GdIN7nOH3ce45WePCcu9MNUugSicyC>

1	marital status/ or divorce/ or marriage/ or single person/ or widowhood/ or spouses/

2	(marital or spousal or romantic relationship* or unmarried or married or partnership* or marriage* or wife or wives or husband* or spouse* or domestic partner* or break-up or divorce* or widow* or spinster* or bachelor* or girlfriend* or boyfriend*).mp.

3	1 or 2

4	exp Myocardial Infarction/

5	exp acute coronary syndrome/

6	exp coronary disease/

7	(myocardial infarction or acute coronary syndrome or coronary heart disease or isch?emic heart disease or heart attack* or AMI).mp.

8	4 or 5 or 6 or 7

9	cerebrovascular disorders/

10	exp stroke/

11	(Stroke* or cerebrovascular disease* or cerebrovascular accident*).mp.

12	9 or 10 or 11

13	8 or 12

14	(HR-PRO or HRPRO or HRQL or HRQoL or QL or QoL).ti,ab. or quality of life.mp. or (health index* or health indices or health profile*).ti,ab. or health status.mp. or ((patient or self or child or parent or carer or proxy) adj (appraisal* or appraised or report or reported or reporting or rated or rating* or based or assessed or assessment*)).ti,ab. or ((disability or function or functional or functions or subjective or utility or utilities or wellbeing or well being) adj2 (index or indices or instrument or instruments or measure or measures or questionnaire* or profile or profiles or scale or scales or score or scores or status or survey or surveys)).ti,ab.

15	exp patient reported outcome measures/

16	exp patient outcome assessment/

17	("patient-reported outcome*" or (((self adj report*) or (patient adj2 (reported or centered or centred or preference* or experience* or perception or perceived))) and (measure* or questionnaire* or survey* or psychometric* or depress* or psychologic* or anxiety or symptom* or fatigue))).mp.

18	15 or 16 or 17

19	(EQ-5D or SF-12 or SF-36 or "general health questionnaire" or PROMIS).mp.

20	(Neuro-QOL or stroke impact scale or stroke specific qol or aphasia quality of life or SAQOL or stroke-adapted sickness impact profile or SA-SIP).mp.

21	(seattle angina questionnaire or SAQ or Mac-New Questionnaire or HeartQoL or QLICD_CHD).mp.

22	19 or 20 or 21

23	14 or 18 or 22

24	3 and 13 and 23

**2.2 Search strategy for Web of Science:**

<https://www.webofscience.com/wos/woscc/summary/2f623710-a68d-4661-aa73-e9e293499af1-1dfe6fc3/relevance/1>

1	TS=("marital status" OR divorce OR marriage OR "single person" OR widowhood OR spouses)

2	TS=(marital OR spousal OR "romantic relationship*" OR unmarried OR married OR partnership* OR marriage* OR wife OR wives OR husband* OR spouse* OR "domestic partner*" OR break-up OR divorce* OR widow* OR spinster* OR bachelor* OR girlfriend* OR boyfriend*)

3	#2 OR #1

4	TS="myocardial infarction"

5	TS="acute coronary syndrome"

6	TS="coronary disease"

7	TS=("ischemic heart disease" OR "ischaemic heart disease" OR "heart attack*" OR AMI)

8	#4 OR #5 OR #6 OR #7

9	TS="cerebrovascular disorders"

10	TS=stroke

11	TS=("cerebrovascular disease*" OR "cerebrovascular accident*")

12	#9 OR #10 OR #11

13	#8 OR #12

14	TI=(HR-PRO or HRPRO or HRQL or HRQoL or QL or QoL or "health index*" or "health indices" or "health profile*") OR AB=(HR-PRO or HRPRO or HRQL or HRQoL or QL or QoL or "health index*" or "health indices" or "health profile*") OR TS=("quality of life" OR "health status") OR TI= ((patient or self or child or parent or carer or proxy) NEAR (appraisal* or appraised or report or reported or reporting or rated or rating* or based or assessed or assessment*)) OR AB= ((patient OR self or child or parent or carer or proxy) NEAR (appraisal* or appraised or report or reported or reporting or rated or rating* or based or assessed or assessment*)) OR TI=((disability or function or functional or functions or subjective or utility or utilities or wellbeing or "well being") NEAR (index or indices or instrument or instruments or measure or measures or questionnaire* or profile or profiles or scale or scales or score or scores or status or survey or surveys)) OR AB=((disability or function or functional or functions or subjective or utility or utilities or wellbeing or "well being") NEAR (index or indices or instrument or instruments or measure or measures or questionnaire* or profile or profiles or scale or scales or score or scores or status or survey or surveys))

15	TS="patient reported outcome measures"

16	TS="patient outcome assessment"

17	TS=("patient-reported outcome*" OR ((patient OR self) AND report* AND (outcome* OR measure* OR questionnaire* OR survey* OR psychometric*)))

18	#15 or #16 or #17

19	ALL=(EQ-5D OR SF-36 OR SF-12 OR "general health questionnaire" OR PROMIS)

20	ALL=(Neuro-QOL OR "stroke impact scale" OR "stroke specific qol" OR "aphasia quality of life" OR SAQOL OR "stroke-adapted sickness impact profile" OR SA-SIP)

21	ALL=("seattle angina questionnaire" OR SAQ OR "Mac-New Questionnaire" OR HeartQoL OR QLICD_CHD)

22	#19 or #20 or #21

23	#22 OR #18

24	#3 AND #13 AND #23

**2.3 Search strategy for Scopus:**

( INDEXTERMS ( "marital status" )  OR  INDEXTERMS ( divorce )  OR  INDEXTERMS ( marriage )  OR  INDEXTERMS ( "single person" )  OR  INDEXTERMS ( widowhood )  OR  INDEXTERMS ( spouses )  OR  TITLE-ABS-KEY ( marital  OR  spousal  OR  "romantic relationship*"  OR  unmarried  OR  married  OR  partnership*  OR  marriage*  OR  wife  OR  wives  OR  husband*  OR  spouse*  OR  "domestic partner*"  OR  break-up  OR  divorce*  OR  widow*  OR  spinster*  OR  bachelor*  OR  girlfriend*  OR  boyfriend* ) )

AND

( INDEXTERMS ( "cerebrovascular disorders" )  OR  INDEXTERMS ( stroke )  OR  TITLE-ABS-KEY ( stroke*  OR  "cerebrovascular disease*"  OR  "cerebrovascular accident*" )  OR  INDEXTERMS ( "Myocardial Infarction" )  OR  INDEXTERMS ( "acute coronary syndrome" )  OR  INDEXTERMS ( "coronary disease" )  OR  TITLE-ABS-KEY ( "myocardial infarction"  OR  "acute coronary syndrome"  OR  "coronary heart disease"  OR  "isch?emic heart disease"  OR  "heart attack*"  OR  ami ) )

AND

ALL ( "patient-reported outcome*" )  OR  TITLE-ABS-KEY ( ( ( self  W/1  report* )  OR  ( patient  W/2  ( reported  OR  centered  OR  centred  OR  preference*  OR  experience*  OR  perception  OR  perceived ) )  AND  ( measure*  OR  questionnaire*  OR  survey*  OR  psychometric*  OR  depress*  OR  psychologic*  OR  anxiety  OR  symptom*  OR  fatigue ) ) )  OR  INDEXTERMS ( "patient reported outcome measures" )  OR  INDEXTERMS ( "patient outcome assessment" )  OR  TITLE-ABS-KEY ( eq-5d  OR  sf-12  OR  sf-36  OR  "general health questionnaire"  OR  promis )  OR  TITLE-ABS-KEY ( neuro-qol  OR  "stroke impact scale"  OR  "stroke specific qol"  OR  "aphasia quality of life"  OR  saqol  OR  "stroke-adapted sickness impact profile"  OR  sa-sip )  OR  TITLE-ABS-KEY ( "seattle angina questionnaire"  OR  saq  OR  "Mac-New Questionnaire"  OR  heartqol  OR  qlicd_chd )  OR  TITLE-ABS ( hr-pro  OR  hrpro  OR  hrql  OR  hrqol  OR  ql  OR  qol )  OR  TITLE-ABS-KEY ( "quality of life" )  OR  TITLE-ABS ( "health index*"  OR  "health indices"  OR  "health profile*" )  OR  TITLE-ABS-KEY ( "health status" )  OR  TITLE-ABS ( ( patient  OR  self  OR  child  OR  parent  OR  carer  OR  proxy )  W/1  ( appraisal*  OR  appraised  OR  report  OR  reported  OR  reporting  OR  rated  OR  rating*  OR  based  OR  assessed  OR  assessment* ) )  OR  TITLE-ABS ( ( disability  OR  function  OR  functional  OR  functions  OR  subjective  OR  utility  OR  utilities  OR  wellbeing  OR  "well being" )  W/2  ( index  OR  indices  OR  instrument  OR  instruments  OR  measure  OR  measures  OR  questionnaire*  OR  profile  OR  profiles  OR  scale  OR  scales  OR  score  OR  scores  OR  status  OR  survey  OR  surveys ) )

**2.4 Search strategy for EMBASE (via Ovid):**

<https://ovidsp.ovid.com/ovidweb.cgi?T=JS&NEWS=N&PAGE=main&SHAREDSEARCHID=6m3DabQezlb6lABOv0CsXCfaE7TzA3O4jHRXh9zorqGIjWt4HBrZiBTkoBzE7RvMk>

1	marriage/ or "single person"/ or exp widowed person/ or spouse/

2	(marital or spousal or "romantic relationship*" or unmarried or married or partnership* or marriage* or wife or wives or husband* or spouse* or "domestic partner*" or break-up or divorce* or widow* or spinster* or bachelor* or girlfriend* or boyfriend*).mp.

3	1 or 2

4	heart Infarction/

5	acute coronary syndrome/

6	coronary artery disease/

7	("myocardial infarction" or "acute coronary syndrome" or "coronary heart disease" or "isch?emic heart disease" or "heart attack*" or AMI).mp.

8	4 or 5 or 6 or 7

9	cerebrovascular disease/

10	(Stroke* or "cerebrovascular disease*" or "cerebrovascular accident*").mp.

11	9 or 10

12	8 or 11

13	(HR-PRO or HRPRO or HRQL or HRQoL or QL or QoL).ti,ab. or quality of life.mp. or (health index* or health indices or health profile*).ti,ab. or health status.mp. or ((patient or self or child or parent or carer or proxy) adj (appraisal* or appraised or report or reported or reporting or rated or rating* or based or assessed or assessment*)).ti,ab. or ((disability or function or functional or functions or subjective or utility or utilities or wellbeing or well being) adj2 (index or indices or instrument or instruments or measure or measures or questionnaire* or profile or profiles or scale or scales or score or scores or status or survey or surveys)).ti,ab.

14	patient-reported outcome/

15	outcome assessment/

16	("patient-reported outcome*" or (((self adj report*) or (patient adj2 (reported or centered or centred or preference* or experience* or perception or perceived))) and (measure* or questionnaire* or survey* or psychometric* or depress* or psychologic* or anxiety or symptom* or fatigue))).mp.

17	14 or 15 or 16

18	(EQ-5D or SF-12 or SF-36 or "general health questionnaire" or PROMIS).mp.

19	(Neuro-QOL or "stroke impact scale" or "stroke specific qol" or "aphasia quality of life" or SAQOL or "stroke-adapted sickness impact profile" or SA-SIP).mp.

20	("seattle angina questionnaire" or SAQ or "Mac-New Questionnaire" or HeartQoL or QLICD_CHD).mp.

21	18 or 19 or 20

22	13 or 17 or 21

23	3 and 12 and 22

**2.5 Search strategy for PsycINFO (via Ovid):**

<https://ovidsp.ovid.com/ovidweb.cgi?T=JS&NEWS=N&PAGE=main&SHAREDSEARCHID=458lNfsG47vrkLAWdtduLnNgo0XPs2NUqpqEIirrJTO2Uohtb8wlOP5WosFvr1Qir>

1	marital status/ or "marital separation"/ or divorce/ or marriage/ or "single persons"/ or widowers/ or widows or exp spouses/ or "marital separation"/ or "significant others"/

2	(marital or spousal or "romantic relationship*" or unmarried or married or partnership* or marriage* or wife or wives or husband* or spouse* or "domestic partner*" or break-up or divorce* or widow* or spinster* or bachelor* or girlfriend* or boyfriend*).mp.

3	1 or 2

4	exp "Myocardial Infarctions"/

5	("myocardial infarction" or "acute coronary syndrome" or "coronary heart disease" or "isch?emic heart disease" or "heart attack*" or AMI).mp.

6	4 or 5

7	"cerebrovascular disorders"/

8	"cerebrovascular accidents"/

9	(Stroke* or "cerebrovascular disease*" or "cerebrovascular accident*").mp.

10	7 or 8 or 9

11	6 or 10

12	(HR-PRO or HRPRO or HRQL or HRQoL or QL or QoL).ti,ab. or quality of life.mp. or (health index* or health indices or health profile*).ti,ab. or health status.mp. or ((patient or self or child or parent or carer or proxy) adj (appraisal* or appraised or report or reported or reporting or rated or rating* or based or assessed or assessment*)).ti,ab. or ((disability or function or functional or functions or subjective or utility or utilities or wellbeing or well being) adj2 (index or indices or instrument or instruments or measure or measures or questionnaire* or profile or profiles or scale or scales or score or scores or status or survey or surveys)).ti,ab.

13	exp "patient reported outcome measures"/

14	("patient-reported outcome*" or (((self adj report*) or (patient adj2 (reported or centered or centred or preference* or experience* or perception or perceived))) and (measure* or questionnaire* or survey* or psychometric* or depress* or psychologic* or anxiety or symptom* or fatigue))).mp.

15	13 or 14

16	(EQ-5D or SF-12 or SF-36 or "general health questionnaire" or PROMIS).mp.

17	(Neuro-QOL or "stroke impact scale" or "stroke specific qol" or "aphasia quality of life" or SAQOL or "stroke-adapted sickness impact profile" or SA-SIP).mp.

18	("seattle angina questionnaire" or SAQ or "Mac-New Questionnaire" or HeartQoL or QLICD_CHD).mp.

19	16 or 17 or 18

20	12 or 15 or 19

21	3 and 11 and 20

**Supplemental Table 3. Data extraction form**

| **Reviewer:** | **Date:** |  |
| --- | --- | --- |
| **Study characteristics** | | |
| Author last name |  |  |
| Publication year | Country |  |
| Study type |  |  |
| ( ) Cohort | ( ) Case control | ( ) Cross sectional |
| Setting | ( ) Hospital-based | ( ) Population-based |
| Condition | ( ) AMI | ( ) Stroke |
| **Participants (individual/proxy/other)** | | |
| Sample size | Age (mean) | Sex: Male (n;%) Female (n;%) |
| Other notes |  |  |
| **Exposure of interest** | | |
| Definition of marital/partner status | ( ) Self-report | ( ) Other |
| Reference group (categories) |  |  |
| Other notes |  |  |
| **Outcomes** |  |  |
| PROM name |  |  |
| Definition |  |  |
| Domain measured |  |  |
| Measure time |  |  |
| Score in each group |  |  |
| Other notes |  |  |
| **Results** |  |  |
| # Participants with outcome |  |  |
| Mean score for each exposure group | | |
| Association type (OR/RR/HR/Other:) | | |
| Unadjusted/Adjusted association |  |  |
| Covariates adjusted |  |  |
| Sex-specific results (if any) |  |  |
| Other notes |  |  |
| **Author’s conclusion:** |  |  |
| **Reviewer comments:** |  |  |
